# Does sensory modulation dysfunction contribute to emotional dysregulation in children with ADHD?: Analysis plan

**DOI:** 10.1101/2020.10.09.20191601

**Authors:** Alisha Bruton, Angela Senders, Brenda Leung, Irene Hatsu, L. Eugene Arnold, Jeanette Johnstone

**Affiliations:** Oregon Health & Science University, Department of Psychiatry, Portland, OR, USA; National University of Natural Medicine, Helfgott Research Institute, Portland, OR, USA; Faculty of Health Sciences, University of Lethbridge, Lethbridge, Alberta, Canada; Department of Human Sciences, The Ohio State University, Columbus, OH, USA; Department of Psychiatry & Behavioral Health, The Ohio State University, Columbus, OH, USA

## Abstract

**Introduction:** Attention-deficit/hyperactivity disorder (ADHD) is one of the most common neurodevelopmental disorders of childhood. Up to 50% of children with ADHD may also experience symptoms of emotional dysregulation, such as anger, irritability, and aggression. Emotional dysregulation contributes to adverse health outcomes such as depression and peer problems, yet it is poorly understood, and effective treatment options are lacking. Emerging evidence suggests that sensory processing may play a role in emotional dysregulation. Forty to 50% of children with ADHD may also experience sensory modulation dysfunction, or SMD. SMD is characterized by hypo- or hyperreactivity to pain and sensation. Only one study investigated the relationship of SMD and emotional dysregulation in ADHD; they found a correlation of r=0.45. If SMD drives emotional dysregulation in ADHD, treating SMD has the potential to improve emotional regulation. Further evaluating the relationship between SMD and emotional dysregulation in ADHD is the crucial first step in developing effective treatment options.

**Methods:** Data for this analysis are derived from the baseline assessment of a multi-site, randomized, controlled trial: The Micronutrients for ADHD in Youth (MADDY) Study. The study enrolled children aged 6-12 with a diagnosis of ADHD and symptoms of emotional dysregulation. Using a cross-sectional study design, we will measure the association between emotional dysregulation and SMD at baseline. Emotional dysregulation was measured using the Strengths and Difficulties Questionnaire (SDQ) and a composite score from the Child and Adolescent Symptom Inventory, Version-5 (CASI-5). SMD will be assessed using two subscales from the Temperament in Middle Childhood Questionnaire (TMCQ). To test our hypothesis, we will use simple linear regression. Models will be adjusted for potential confounding variables.

**Conclusion:** Our results will serve to better characterize the relationship between SMD and emotional dysregulation in children with ADHD, which may inform treatment options and diminish adverse health outcomes.

## BACKGROUND

Attention-deficit/hyperactivity disorder (ADHD) is one of the most common neurodevelopmental disorders of childhood.^1^ It is defined by behavioral and cognitive symptoms such as hyperactivity, impulsivity, forgetfulness, and inattention.^2^ Up to 50% of children with ADHD may also experience emotional symptoms such as anger, irritability, or aggression, collectively called emotional dysregulation.^3,4^ Emotional dysregulation and ADHD are closely linked genetically, which may explain their frequent comorbidity.^5^ Emotional dysregulation is associated with negative outcomes such as depression, peer problems, and expulsion from school, yet the condition remains poorly understood and effective treatment options are lacking.^6–8^ Emotional dysregulation manifests in multiple mood and psychiatric disorders, thus identifying effective treatments has important trans-diagnostic implications.^9–11^

Emerging evidence suggests that sensory processing may play a role in emotional dysregulation. Sensory modulation dysfunction (SMD) is the inability to respond to environmental stimuli in a way appropriate to the magnitude of the stimulus, and is characterized by hyper- or hyposensitivity to sensation and pain.^12–16^ SMD is associated with emotional dysregulation in cross-sectional studies of children with behavioral problems, with *r* values ranging from 0.31-0.83.^13,16^ Forty to 50% of children with ADHD may also experience SMD.^12,17^ Only one study has investigated the association between emotional regulation and SMD in children with ADHD; they found an association of *r=*0.45.^15^ This is problematic gap in the research. If SMD drives emotional dysregulation in ADHD, treating SMD has the potential to improve emotional regulation which may prevent or reduce many of the negative personal, social, and academic outcomes which can occur without effective management. Further evaluating the relationship between SMD and emotional dysregulation in ADHD is the crucial first step in developing effective treatment options.

Therefore, we propose to investigate the relationship between SMD and emotional dysregulation in children with ADHD. We are uniquely situated to address this question using data from the Micronutrients for ADHD Youth (MADDY) Study.

### MADDY study background

We propose to examine SMD in a cross-sectional sample of children with symptoms of ADHD and emotional dysregulation. The questionnaire data were collected as part of a 3-site randomized controlled trial (RCT) called the Micronutrients in ADHD Youth (MADDY) study. The three trial sites include Oregon Health & Science University (OHSU), Ohio State University (OSU), and University of Lethbridge (Alberta, Canada). Each site enrolled 45-50 participants for a total of n=145.

The study consisted of an 8-week intervention during which participants were randomly assigned to receive a micronutrient supplement or a placebo, in a ratio of 3:2, respectively. This was followed by an 8-week open-label follow-up, during which all participants were given the opportunity to take the micronutrient supplement. Questionnaire data were collected every 4 weeks, and biological samples, including blood, saliva, urine, hair, and stool, were collected baseline and at week 8; hair, saliva, and stool were collected at week 16. For this project, we will examine questionnaire data from the baseline visit only.

The main study protocol was approved by the institutional review board at OHSU and is detailed elsewhere.^18^ For the sake of transparency, the protocol for this proposed project will be written in advance and indexed on the MedRxiv platform before the analysis begins.^19^ This is an update to an earlier analysis plan indexed on MedRxiv. Our original plan was based on a factor analysis of the TMCQ which was in progress and has since been completed. The final factor structure does not include one of our proposed independent variables, sensation seeking, thus this has been dropped.

Our research question is: At baseline, is SMD associated with emotional dysregulation among children with ADHD? SMD will be measured with the Temperament in Middle Childhood Questionnaire (TMCQ). Emotional dysregulation will be measured with the Strengths and Difficulties Questionnaire (SDQ) and specific subscales from the Child and Adolescent Symptom Inventory, version 5 (CASI-5); the Inattention, Hyperactivity, Oppositional Defiant Disorder (ODD), Peer Conflict, and Disruptive Mood Dysregulation Disorder (DMDD) subscales.

We hypothesize that there is a positive association between SMD and emotional dysregulation in this population, that is, individuals with higher levels of SMD will have higher levels of emotional dysregulation, as measured by the SDQ and CASI-5 questionnaires.

## METHODS

### Study Sample

#### Inclusion criteria

Children ages 6-12 who completed the baseline visit of the MADDY study.

#### Exclusion criteria

Children missing baseline assessments of dependent or independent variables.

### Variables

#### Primary independent variable: Sensory Modulation Dysfunction (SMD)

We will examine aspects of SMD using two variables, each of which are derived from the TMCQ. MADDY participants completed the TMCQ at baseline. The TMCQ is a validated, 157-item, parent-report questionnaire used to assess dimensions of childhood temperament. Each question has five Likert-type answers for which the parent indicated the accuracy of a statement about the child’s character or behavior. A recent factor analysis of the TMCQ (Antovich, Nigg, manuscript in preparation) identified two subscales that measure aspects of SMD, including pain sensitivity and perceptual sensitivity. The subscales consist of 7 and 10 questions, respectively. The total score for each subscale is a mean of each of the individual questions, and ranges from 1-5. A higher score indicates higher levels of the construct being measured. We will evaluate each of the two subscales as a continuous variable.

#### Dependent variable: Emotional dysregulation

We will evaluate emotional dysregulation using three variables. Our primary dependent variables are two subscales from the SDQ: the emotional problems subscale and the conduct problems subscale. The SDQ is a validated, 25-item, parent-report questionnaire used to assess emotional and behavioral problems in children. It includes five subscales, each of which has five questions. Each question has three possible answers, which indicate the accuracy of a statement about the child’s behavior or emotional state. Answers include not true, somewhat true, and certainly true, coded in a Likert scale of 0-2. Subscale scores are obtained by adding the answers from each individual question in that subscale. Possible subscale scores range from 0-10, and a higher score indicates more of the symptom or quality in question. We chose these subscales because they address emotional symptoms including unhappiness, nervousness, fear, worrying, and temper tantrums. They have been used as outcome measures in other studies of children with ADHD and emotional dysregulation.^20^ We will evaluate each of the two subscales as a continuous variable.

Additionally, we will evaluate the CASI-5 composite score as a secondary dependent variable. The CASI-5 is a validated, parent-report questionnaire used to assess psychiatric and behavioral symptoms in children. It is comprised of subscales which correspond to DSM-5 diagnostic criteria for specific conditions. Each subscale has questions that have four possible answers. The answers indicate the frequency with which the child experiences particular symptoms: never, sometimes, often, and very often, using a Likert scale of 0-3. Higher scores indicate more of the construct being measured. Each subscale also includes a single question about the impairment caused by those symptoms. Composite scores are designed to be a more robust measure of emotional dysregulation than single subscales alone.^21^ This composite scoring method was developed to assess symptoms of emotional dysregulation in children with ADHD, as many questionnaires designed to measure ADHD symptoms do not accurately capture emotional symptoms.^21^ We will use the equal subscales weighting method, which combines the Inattention, Hyperactivity, Oppositional Defiant Disorder (ODD), Peer Conflict, and Disruptive Mood Dysregulation Disorder (DMDD) subscales. It also incorporates the impairment caused by the symptoms in each of those categories. Possible scores vary from 0-3. It will be evaluated as a continuous variable.

#### Potential confounders and other independent variables

The following variables will be assessed for potential confounding or addition to the model: age, gender, race, study site, and season of enrollment. Race will be included as a proxy for racism and other social stressors that participants may be exposed to.^22^ Multi-site trials may have procedural differences between sites; therefore, we will assess study site as an additional independent variable. Because the school or home environment may impact behavioral or emotional symptoms, we will also assess season of enrollment to account for differences in the time of year at which the baseline visit occurred.

### Analysis plan

#### Descriptive analysis

Statistical analyses will be performed with the Stata software, version 16.1.^23^ Code will be cataloged and made available on the GitHub platform. Baseline characteristics such as age, gender, race, season of enrollment, and study site will be examined using descriptive statistics, and will be presented as a number (n) and percentage (%), or mean +/- SD. Because our primary independent variables are continuous, we will divide them into quartiles and present the baseline characteristics across those quartiles (Mock Table 1). We will visualize the relationship between the TMCQ subscales and the SDQ and CASI-5 composite score using scatter plots and a correlation coefficient for each association. Descriptive analyses will inform subsequent hypothesis testing, and the STROBE guidelines will be used to guide reporting.^24^

**Mock Table 1:**
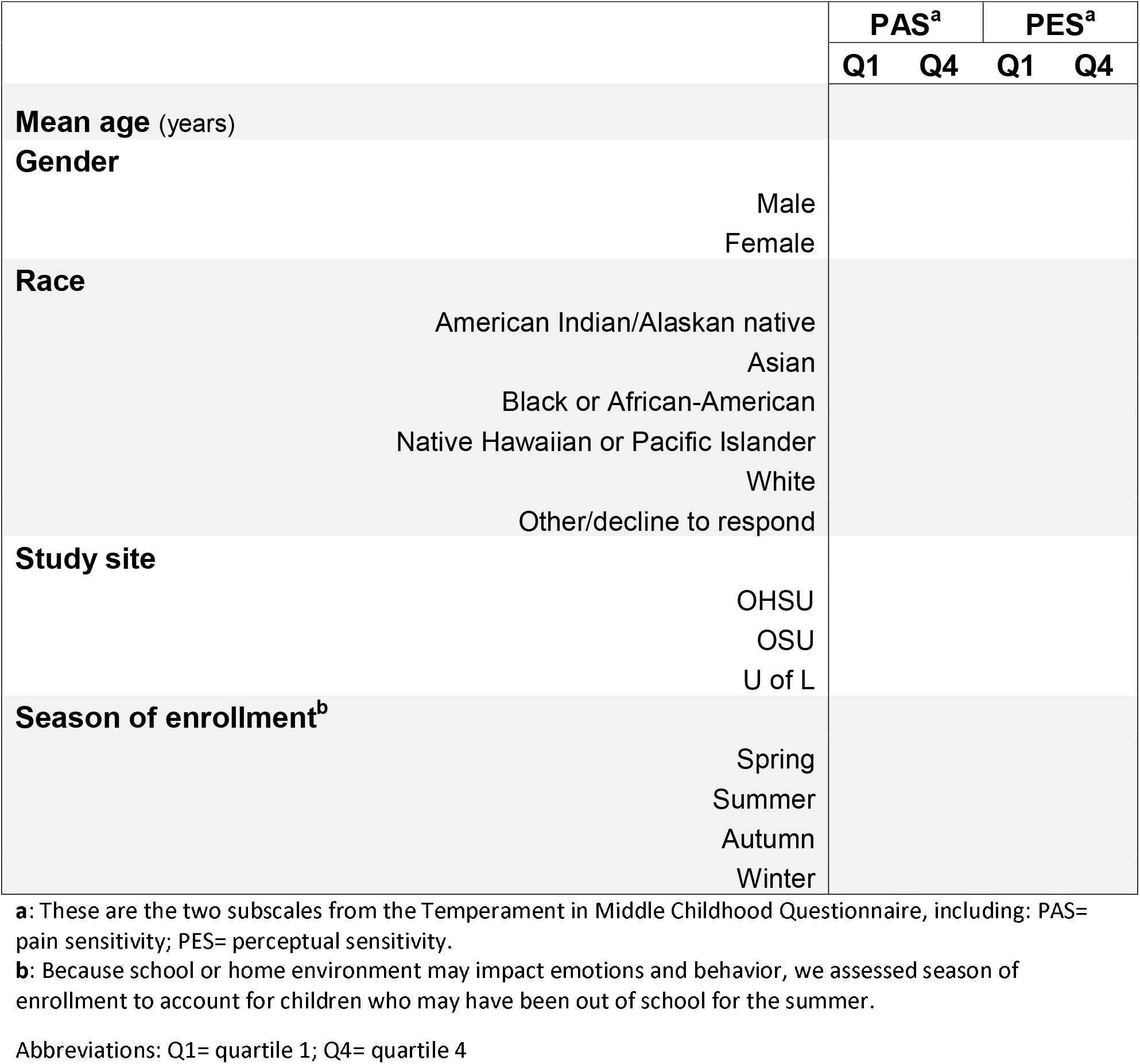
Demographic characteristics of participants by quartile of each primary independent variable (pain sensitivity, perceptual sensitivity)

#### Multivariable analyses

We hypothesize that SDQ subscales and CASI-5 composite scores will be higher among participants with higher scores on the subscales of the TMCQ. Therefore, the TMCQ subscales are specified as two separate independent variables and the two SDQ subscales and CASI-5 composite score will be modeled as three separate dependent variables.

Our six linear models include the following:

- Pain sensation regressed on:
  - Emotional problems (SDQ)
  - Conduct problems (SDQ)
  - Composite score (CASI-5)
- Perceptual sensation regressed on:
  - Emotional problems (SDQ)
  - Conduct problems (SDQ)
  - Composite score (CASI-5)

To test our hypotheses, we will use multiple linear regression. Each of the two subscales of the TMCQ (pain sensitivity and perceptual sensitivity) will be regressed separately on the three dependent variables, for a total of six models. For each model, the unstandardized *β* coefficient and 95% confidence interval (CI) will be estimated as the measure of association between the TMCQ subscale and the SDQ subscales and CASI-5 composite score (Mock Tables 2-3). The *β* coefficient represents the estimated difference in emotional dysregulation scores for every one-point increase in either pain sensitivity or perceptual sensitivity. All hypothesis tests will be two-tailed, and a value of α<0.05 will be used to indicate significance.

**Mock Table 2:**
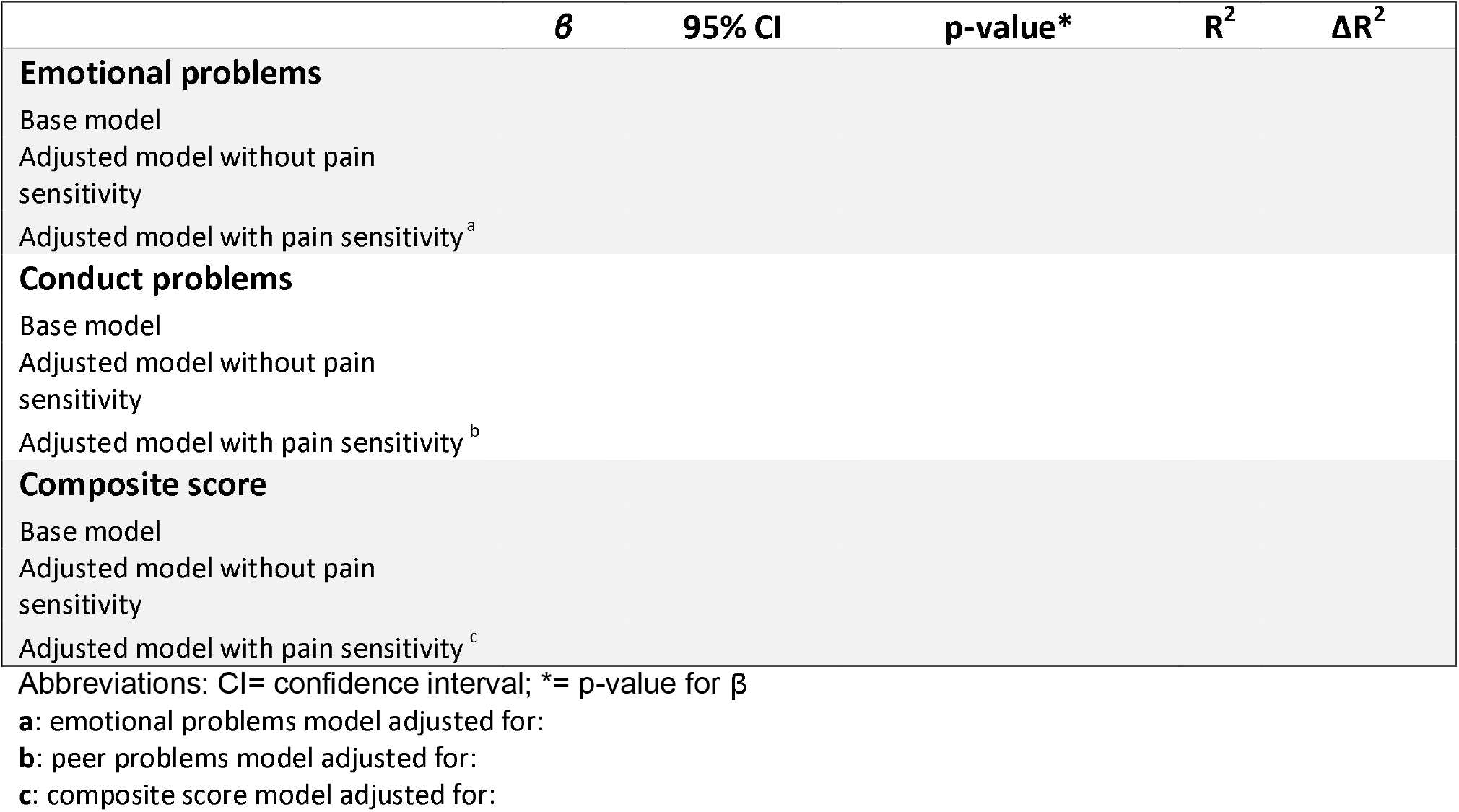
Baseline association (β, 95% CI, R^2^) of pain sensitivity and emotional dysregulation

**Mock Table 3:**
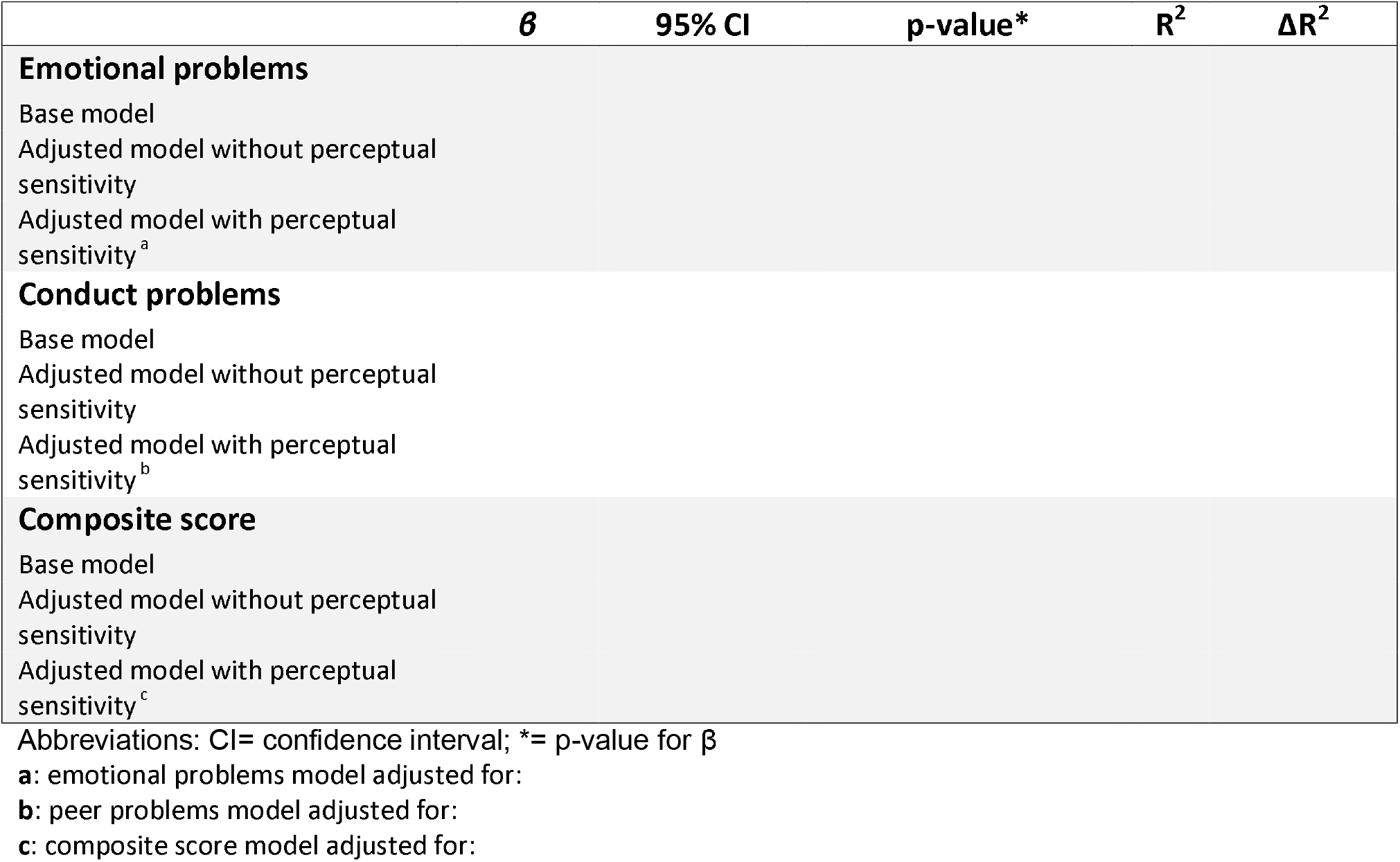
Baseline association (β, 95% CI, R^2^) of perceptual sensitivity and emotional dysregulation

#### Control for confounding

The following methods to control for confounding will be applied to all linear models.

All six base models will be adjusted for age, gender, and study site. Additional variables including race and season of enrollment will be assessed for addition to the final model. Selection of variables to include in the final model will be based on a combination of causal modeling drawn from knowledge of known associations between variables which have been established in the literature and statistical association. Factors which have been reliably demonstrated to cause the exposure, the outcome, or both, will be included each of the models, regardless of statistical significance. Theoretical or unestablished confounders will be selected based on the 10% change in estimate method.^25^ They will be assessed statistically by adding each variable to the base model one at a time. Any variable that results in ≥10% change in the *β* coefficient when added to the base model will be considered a confounder. After statistical confounders are identified, they will be ranked according to the strength of the confounding, added to the model sequentially by rank, and retained if they remain a confounder in the presence of other variables already in the model. This process will be repeated separately for each of the six models.

#### Effect modification

If we observe an association between SMD and emotional dysregulation, we will assess whether any of these associations vary by sex status. We will stratify our linear regression models by sex and qualitatively examine the *β* coefficients for males and females. We will also check for formal interactions between sex and SMD by including a cross-product term of these variables in the full model. If it is significant, we will present models for males and females separately.

#### Model diagnostics and goodness of fit

Linear regression assumes that error terms are normally distributed, mutually independent, homoscedastic, and have a mean value of zero. We will assess data for violation of these assumptions using statistical and graphical methods. Statistical methods will be reported by name, along with cut-off criteria, and graphical methods will be described and made available in a supplemental table. Statistical methods will include the Shapiro-Wilks W test for normality of residuals as well as the Breusch-Pagan test and White’s test for homoscedasticity. We will examine scatter plots of each primary independent and dependent variable for linearity and the presence of outliers. For the linear regression models, normal probability plots and Q-Q plots will be examined for normality and homoscedasticity. If we find evidence of collinearity, this will be managed by centering variables if appropriate or otherwise excluding them from the model.

#### Missing data, potential selection bias

All analyses will be conducted on complete cases. We expect minimal missing data and do not expect a complete case analysis to induce selection bias. Nevertheless, we will assess for any differences between those who are missing and those who are not missing data. If we find meaningful differences, we will explore ways to impute missing variables.

#### Sensitivity analysis

MADDY participants were selected for symptoms of both ADHD and emotional dysregulation. The presence of ADHD symptoms in all participants may contribute to increased composite scores in the entire sample and may decrease our ability to detect meaningful differences in emotional dysregulation. Therefore, we will conduct a sensitivity analysis using the CASI-5 composite score but excluding the subscales on ADHD symptoms (the Inattention and Hyperactivity subscales). We will evaluate this modified composite score in separate linear models with each of our primary independent variables, pain sensitivity and perceptual sensitivity. Assessing for confounding and effect modification will follow the processes outlined above.

#### Power calculation: linear regression models

Our sample size is predetermined by the number of complete baseline observations in the MADDY study (n=125). Therefore, we conducted a power analysis to determine the minimum amount of variance in the dependent variable that could be attributed to the primary independent variable in our models. R^2^ is a statistical measure that represents the percentage of the variability of the dependent variable that is explained by the linear regression model. Contrasting the R^2^ from a fully adjusted model with the R^2^ derived from the same model minus the primary independent variable provides a value of ΔR^2^. Thus, ΔR^2^ represents the amount of variability in the dependent variable that is explained by the independent variable, conditional on the covariates in the model.

We conducted several power calculations for this analysis using the Power Analysis Software (PASS). We varied the correlation between independent variables from 0.10 to 0.40 and found little material difference between the minimally detectable ΔR^2^, therefore we present the power analysis where independent variables are correlated with value of 0.20. Given seven covariates and α=0.05, a sample size of 125 achieves 90% power to detect an ΔR^2^ of at least 6% attributable to our primary independent variable. Literature with regression analyses of sensory modulation dysfunction on emotional regulation is sparse. One study conducted this type of regression analysis in children without ADHD. Their independent variables included sensation seeking, sensory sensitivity, sensory avoidance, and sensory registration, which are similar to our independent variables of pain sensitivity and perceptual sensitivity. They reported crude R^2^ values of 0.24-0.36, therefore we are most likely powered to detect the relationship, if it exists.^13^

## CONCLUSION

Emotional dysregulation in ADHD is impairing yet poorly understood. Sensory modulation dysfunction may contribute to emotional symptoms in ADHD. Investigating this relationship may inform future interventions which can mitigate the negative outcomes associated with emotional dysregulation in children with ADHD.

## Data Availability

Data and code for this project will be made available on GitHub after the project is completed.

https://github.com/alishabruton/TMCQproject

